# A novel method combining mechanical contraction and electrical activation to recommend left ventricular lead placement to improve CRT response

**DOI:** 10.1101/2023.11.18.23298724

**Authors:** Hongjin Si, Zhuo He, Saurabh Malhotra, Xinwei Zhang, Fengwei Zou, Siyuan Xue, Zhiyong Qian, Yao Wang, Xiaofeng Hou, Weihua Zhou, Jiangang Zou

**Author notes:** Hongjin Si and Zhuo He contributed equally to this work. Corresponding authors: Jiangang Zou, MD, PhD, Address: Guangzhou Road 300, Nanjing, Jiangsu, China 210029 or, Weihua Zhou, PhD, Address: 1400 Townsend Drive, Houghton, MI, USA 49931.

## Abstract

**Background:** The segment of the latest mechanical contraction (LMC) does not always overlap with the site of the latest electrical activation (LEA). By integrating both mechanical and electrical dyssynchrony, this study aimed to propose a new method for recommending left ventricular (LV) lead placements, with the goal of enhancing response to cardiac resynchronization therapy (CRT).

**Methods:** The LMC segment was determined by single-photon emission computed tomography myocardial perfusion imaging (SPECT MPI) phase analysis. The LEA site was detected by vectorcardiogram (VCG). The recommended segments for LV lead placement were as follows: (1) the LMC viable segments that overlapped with the LEA site;(2) the LMC viable segments adjacent to the LEA site;(3) If no segment met either of the above, the LV lateral wall was recommended. The response was defined as ≥15% reduction in left ventricular end-systolic volume (LVESV) 6-months after CRT. Patients with LV lead located in the recommended site were assigned to the recommended group, and those located in the non-recommended site were assigned to the non-recommended group.

**Results:** The cohort comprised of 76 patients, including 54 (71.1%) in the recommended group and 22 (28.9%) in the non-recommended group. Among the recommended group, 74.1% of the patients responded to CRT, while 36.4% in the non-recommended group were responders (*p*=.002). Compared to pacing at the non-recommended segments, pacing at the recommended segments showed an independent association with an increased response by univariate and multivariate analysis (odds ratio 5.000, 95% confidence interval 1.731-14.441, *p=*.003; odds ratio 7.326, 95% confidence interval 1.527-35.144, *p=*.013).

**Conclusions:** Our findings indicate that pacing at the recommended segments, by integrating of mechanical and electrical dyssynchrony, is significantly associated with an improved CRT response.

## INTRODUCTION

Cardiac resynchronization therapy (CRT) has been shown to improve clinical outcomes and reverse left ventricular (LV) remodeling in patients with medically refractory heart failure (HF) with LV systolic impairment and electrical dyssynchrony.^1^ However, a significant percentage of patients do not show benefit from CRT.^2^ Notably, patients with left bundle branch block (LBBB) morphology and QRS duration(QRSd) ≥150 ms tend to have better response rates.^3^ The optimal positioning of the LV lead holds fundamental significance in ensuring the effectiveness of CRT.^4^

Numerous studies have been conducted to find the optimal LV lead location. Some early CRT studies advocated for LV lead placement along the LV free wall,^1^ while other demonstrated comparable benefits with leads placed along the anterior, lateral, or posterior wall.^5^ The placement of a LV lead according to purely anatomically defined regions is controversial. Several studies had suggested that positioning the LV lead away from scar and in a non-apical location may provide the greatest benefit.^5,6^

Left ventricular mechanical dyssynchrony (LVMD) plays a crucial role in CRT response.^7^ Single-photon emission computed tomography myocardial perfusion imaging (SPECT MPI) phase analysis can be used to measure LV mechanical dyssynchrony. The SPECT Guided LV Lead Placement for Incremental Benefits to CRT Efficacy (GUIDE-CRT) study conducted by our team found that assessment of the LV latest mechanical contracting (LMC) segment using SPECT images significantly increased the rate of target implantation of LV lead, resulting in incremental improvement in CRT efficacy.^8^ Another study from our group,also confirmed that the placement of a LV lead at or near the site of the LMC has an association with improved response to CRT and long-term prognosis.^9^

In addition to mechanical dyssynchrony, LV electrical dyssynchrony is also an important factor affecting CRT response.^3,10^ It has been demonstrated that positioning the LV lead at the site of latest electrical activation (LEA) was associated with superior clinical outcome.^11^ Rad et al^12^ found that the QRS area measured by vectorcardiogram (VCG) can identify delayed LV lateral wall activation, which performed better than QRS duration and LBBB morphology. VCG is a vector loop of cardiac electrical activity in three-dimensional space, which contains the instantaneous amplitude and direction of each time point in the cardiac cycle.^13,14^

Nevertheless, our previous study has found that the segment of LMC does not always overlap with the LEA site.^15^ To bridge this gap, this study aims to investigate whether the combination of both LMC and LEA could recommend LV lead positions to enhance volumetric response to CRT.

## METHODS

### Study Population

A total of 76 patients with systolic HF were retrospectively enrolled in this study at The First Affiliated Hospital of Nanjing Medical University from October 2011 to October 2021. All the patients were in sinus rhythm with intraventricular conduction delay (QRS duration ≥120 ms), LV ejection fraction (LVEF) ≤35%, New York Heart Association (NYHA) functional class II to IV symptoms, and optimal medical therapy at least 3 months before CRT.

All the patients underwent resting gated SPECT MPI, echocardiography, standard 12-lead electrocardiogram (ECG), and NYHA function classification at baseline. The actual LV lead position was identified according to pre-implantation coronary venous angiograms and post-implantation LV lead fluoroscopy images. Echocardiography and NYHA function classification were reevaluated 6 months after CRT. A clinical response to CRT was defined as a ≥1 NYHA class improvement at 6 months.^16^ The study was approved by the institutional review board, and written informed consent was obtained from all patients.

### Evaluation of LV Function by Echocardiography

Transthoracic echocardiography was performed on all patients by an ultrasound specialist who was blinded to the study before and 6 months after CRT. LV end-systolic volume (LVESV), LV end-diastolic volume (LVEDV), and LVEF were recorded using the 2-dimensional modified biplane Simpson method.^9^ Decrease of LVESV ≥15% on follow-up echocardiography was considered as volumetric response to CRT.^6^

### Evaluation of Segments with the Latest Mechanical Contraction from SPECT MPI

We performed resting ECG-gated SPECT MPI before CRT implantation, following a protocol similar to a previously employed method.^9^ 60-90 minutes after 25-30 mCi of Tc-99m sestamibi injection, SPECT was performed on a dual-headed camera (CardioMD, Philips Medical Systems) with a standard resting protocol. Image reconstruction and reorientation were completed by Emory Reconstruction Toolbox (ERToolbox; Atlanta, GA).

We reoriented the gated and ungated transaxial images into short-axis images, and submitted these results to the phase analysis to measure LVMD^17^ and segments of LMC^18^ and to assess regional myocardial perfusion. ^19^ Myocardial scar was defined as LV samples with <50% of the maximum uptake and a scarred segment was identified as the segment where over 50% of LV samples in the region was myocardial scar. The percentage of tracer uptake was displayed on polar map using a 17-segment model and mean phase angle of each segment was displayed on the 17-segment polar map, like described in our previous study.^9^ Excluding 5 apical segments, septal segments, and scarred segments, the LMC segments were defined as the top 4 phase angles which were within 10 degrees of the maximum phase angle. To simplify, the basal and mid anterolateral, lateral, posterolateral segments were grouped together as the lateral wall. The basal anterior and mid anterior segments were grouped together as the anterior wall. The basal inferior and mid inferior segments were grouped together as the inferior wall. Phase standard deviation (PSD, unit: degree) and 95% bandwidth of phase histogram (PHB, unit: degree) were used to assess global LVMD.

### Evaluation of the LV Site with the Latest Electrical Activation by VCG

A standard 12-lead ECG was recorded at baseline. The digital PDF/XML ECG files with vector graphics were used to extract the original digital ECG-signal. We developed a tool implemented in Python that semi-automatically synthesizes digital ECG signals into a VCG. The Kors transformation matrix was used to transform the 12-lead ECG to VCG.^20^ The direction of the maximal T vector was considered as the LEA site.^14^ Excluding LV apex and septal segments, one of the three candidate sites (anterior, lateral, and inferior) from VCG was recommended (Figure 1).

**Figure 1.**
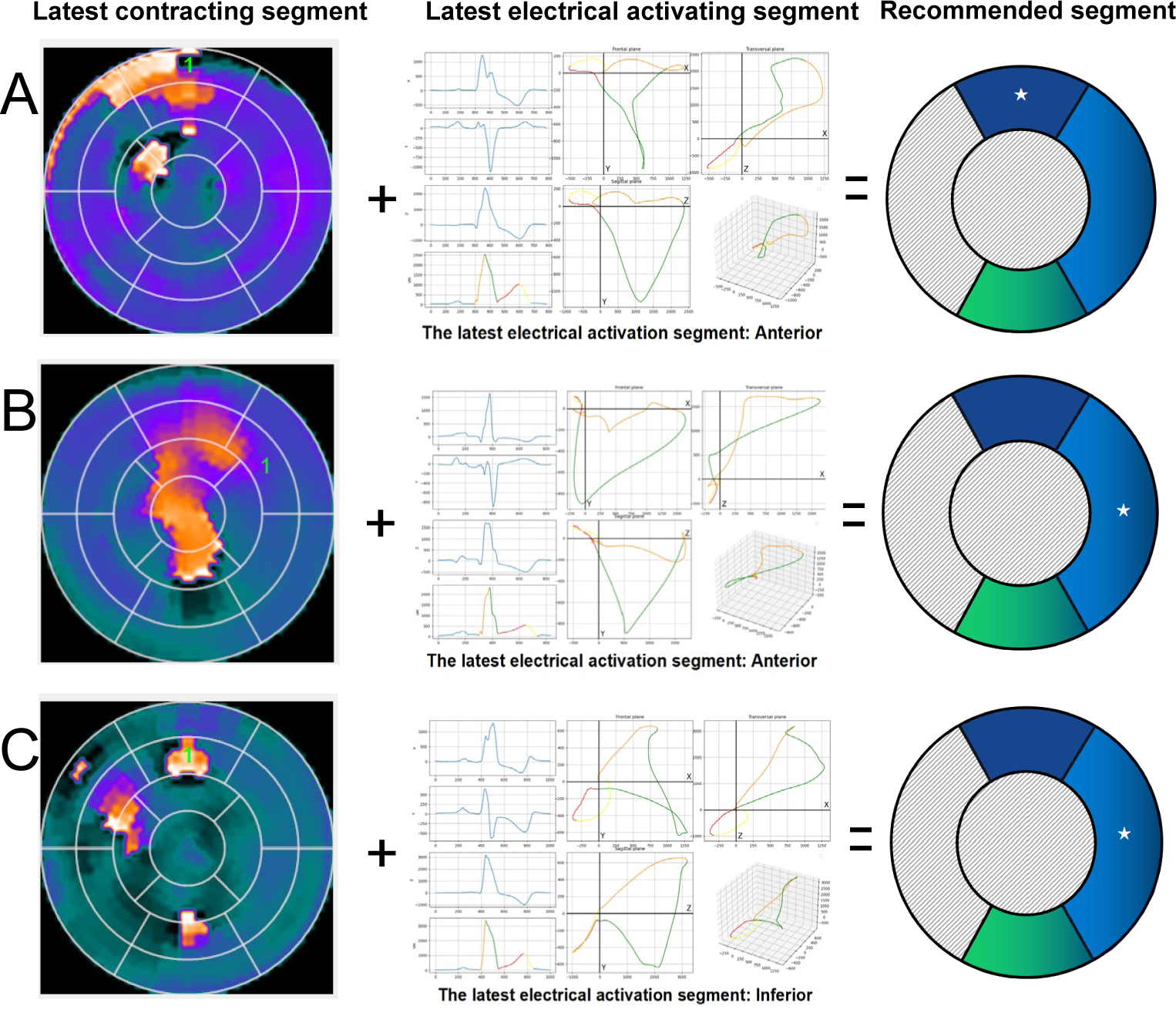
Patient examples of how to recommend the left ventricular lead placement. **(A)** The LMC segment was in the anterior wall and overlapped with the LEA site, so the anterior wall was recommended. **(B)** LMC segment was in the lateral wall and LEA site in the anterior wall, so the lateral wall was recommended. **(C)** LMC segment was in the anterior wall and LEA site in the inferior wall, so the lateral wall was recommended.

### Recommendation of the LV Lead placement segment

After excluding septal segments, LV apex, and segments with more than 50% scar, the segments for LV lead placement were recommended as follows: (1) the LMC viable segments that overlapped with the LEA site(Figure 1A);(2) the LMC viable segments adjacent to the LEA site(Figure 1B);(3) If no segment met either of the above, the LV lateral wall was recommended(Figure 1C).

Patients with LV lead located in the recommended site were assigned to the recommended group, and those located in the non-recommended site were assigned to the non-recommended group.

### Cardiac Resynchronization Therapy implantation

All patients were implanted with a biventricular pacemaker according to standard procedures.^21^ The LV lead was put into the coronary sinus (CS) system and stably positioned into the lateral, posterior, anterolateral, or posterolateral coronary vein by experienced electrophysiologists. The atrial lead was implanted in the right atrial appendage, and the right ventricular (RV) lead was placed at the RV apex or septum.

### Identification of the Actual LV Lead Position

A method similar to the method used in the Multicenter Automatic Defibrillator Implantation Trial–Cardiac Resynchronization Therapy (MADIT-CRT) study^5^ was used for identification of the actual LV lead position. A preimplantation coronary venous angiogram in left anterior oblique 20° to 45° was performed for every patient, and post-implantation fluoroscopic images or CT venography were stored. The left anterior oblique view was used to divide the LV wall into 4 equal parts: anterior, anterolateral, posterolateral, and posterior along the short axis of the heart. To be consistent with SPECT and VCG segmentation, the anterolateral and posterolateral segments were included in the lateral segments. The final LV lead position was categorized as anterior, lateral, or posterior.

### Statistical Analysis

Continuous variables were expressed as mean ± standard deviation (SD). Categorical data are summarized as frequencies and percentages. Comparisons between groups were performed by student’s t-test for parametric variable, Chi-square or Fisher exact tests for categorical variables. The binary logistic regression (*P* <.05 in the univariate regression were included and variables with *P* >.05 in the multivariate regression were excluded) were performed to assess the independent predictors of volumetric response to CRT. Significance was defined as *P* <.05 using two-tailed analysis. Statistical analysis was performed with IBM SPSS Statistics (IBM, Chicago, IL, USA) version 25.0.

## RESULTS

### Patient Characteristics

The baseline characteristics of the 76 enrolled patients are summarized in Table 1. Patients had severely depressed LV function, with a mean LVEF of 27.0±5.0%. Mean LVEDV was 286.3±88.8 ml, and mean LVESV was 211.5±77.3 ml. Forty-one patients (53.9%) had a LV lead position concordant with the LMC segment from SPECT, and 13 patients (17.1%) had a LV lead position concordant with the LEA site by VCG. Fifty-four patients (71.1%) had a recommended LV lead position, whereas 22 (28.9%) had an LV lead positioned in the non-recommended segment. There were no significant differences in baseline data between the two groups, except that, in the recommended group, less patients had diabetes, more patients had a LV lead position concordant with the LMC segment, and more patients had a LV lead in the lateral wall.

**Table 1.**
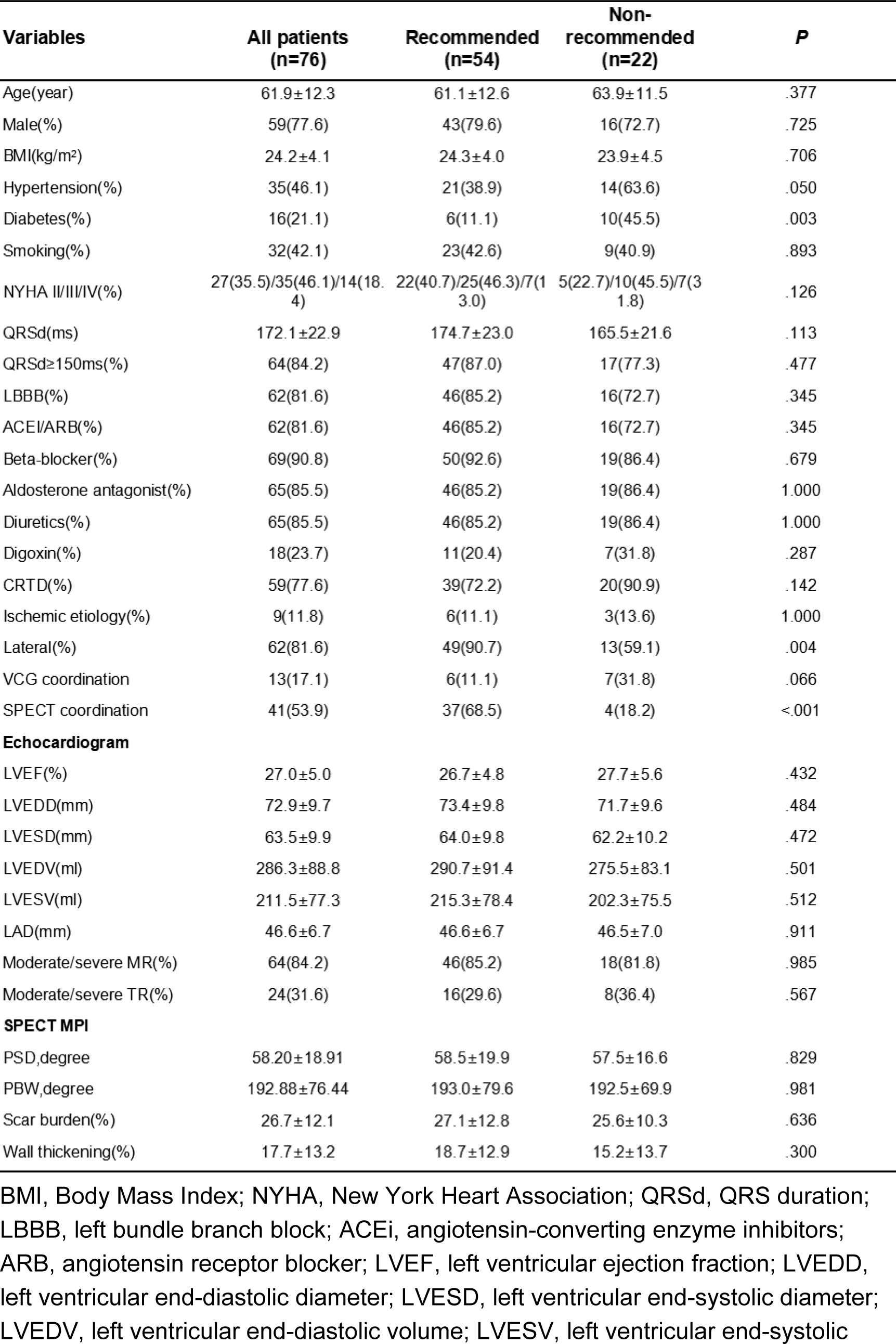
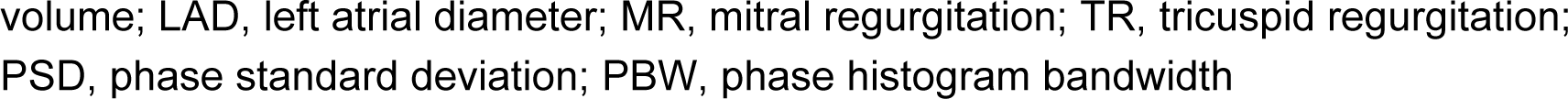
Baseline characteristics of patients in the recommended and non-recommended group BMI, Body Mass Index; NYHA, New York Heart Association; QRSd, QRS duration; LBBB, left bundle branch block; ACEi, angiotensin-converting enzyme inhibitors; ARB, angiotensin receptor blocker; LVEF, left ventricular ejection fraction; LVEDD, left ventricular end-diastolic diameter; LVESD, left ventricular end-systolic diameter; LVEDV, left ventricular end-diastolic volume; LVESV, left ventricular end-systolic volume; LAD, left atrial diameter; MR, mitral regurgitation; TR, tricuspid regurgitation; PSD, phase standard deviation; PBW, phase histogram bandwidth

### Echocardiographic Measurements at Follow-Up

At 6-month follow-up, echocardiographic parameters of patients in the recommended group improved significantly; LVEDV decreased from 290.7±91.4 ml to 219.8±106.2 ml, LVESV from 215.3±78.4 ml to 142.3±91.9 ml, and LVEF increased from 26.7±4.8% to 39.5±12.6% (all *p*<.001). However, patients in the non-recommended group did not have a significant reduction in LV volumes (LVEDV from 275.5±83.1 ml to 245.0±101.8 ml and LVESV from 202.3±75.5 ml to 173.1±91.9 ml, both *p* = NS) and no improvement in LVEF was observed (from 27.7±5.6% to 33.2±11.6%, *p* = NS). The relative change in LVEDV, LVESV, and LVEF showed significant difference between the recommended group and non-recommended group (25.5±19.6% vs. 13.3±20.1%, *p*= .017; 36.6±24.7% vs. 18.0±27.5%, *p*= .005; 48.4±40.6% vs. 18.5±28.0%, *p*= .002) (Figure 2).

**Figure 2.**
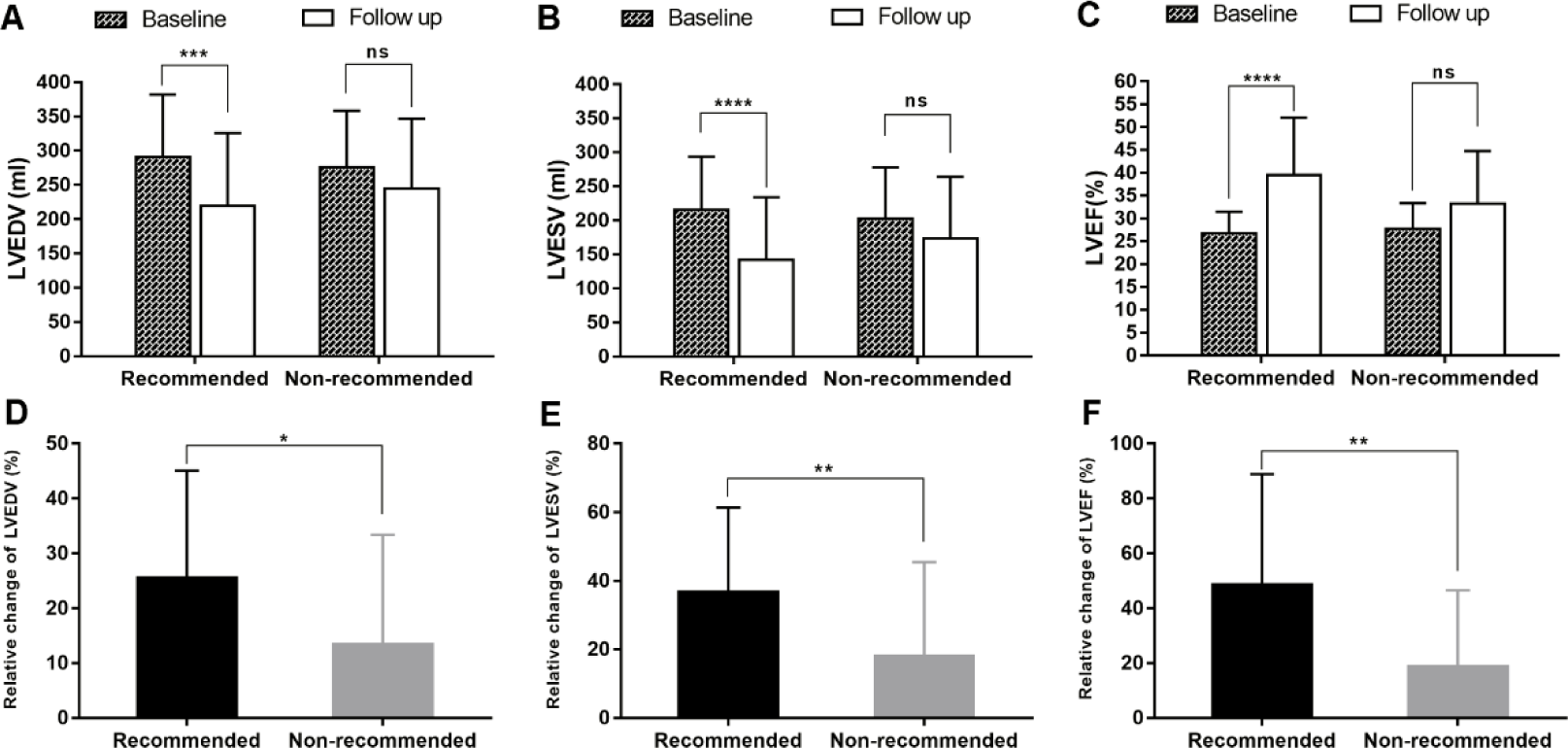
Comparison of echocardiographic measurements between recommended group and non-recommended group. **(A-B-C**): LVEDV, LVESV and LVEF change between baseline and follow up. **(D-E-F):** The relative change of LVEDV, LVESV and LVEF between the recommended and non-recommended group. \**P* < 0.05; \*\**P* < 0.01; \*\*\**P* < 0.001; \*\*\*\**P* < 0.0001; ns, no statistically significant.

### Volumetric Response and Clinical Response

#### Volumetric Response

Six months after CRT implantation, 74.1% (40 of 54) of the patients in the recommended group were responders, while 36.4% (8 of 22) of the patients in the non-recommended group were responders (*p* = .002) (Figure 3A).

**Figure 3.**
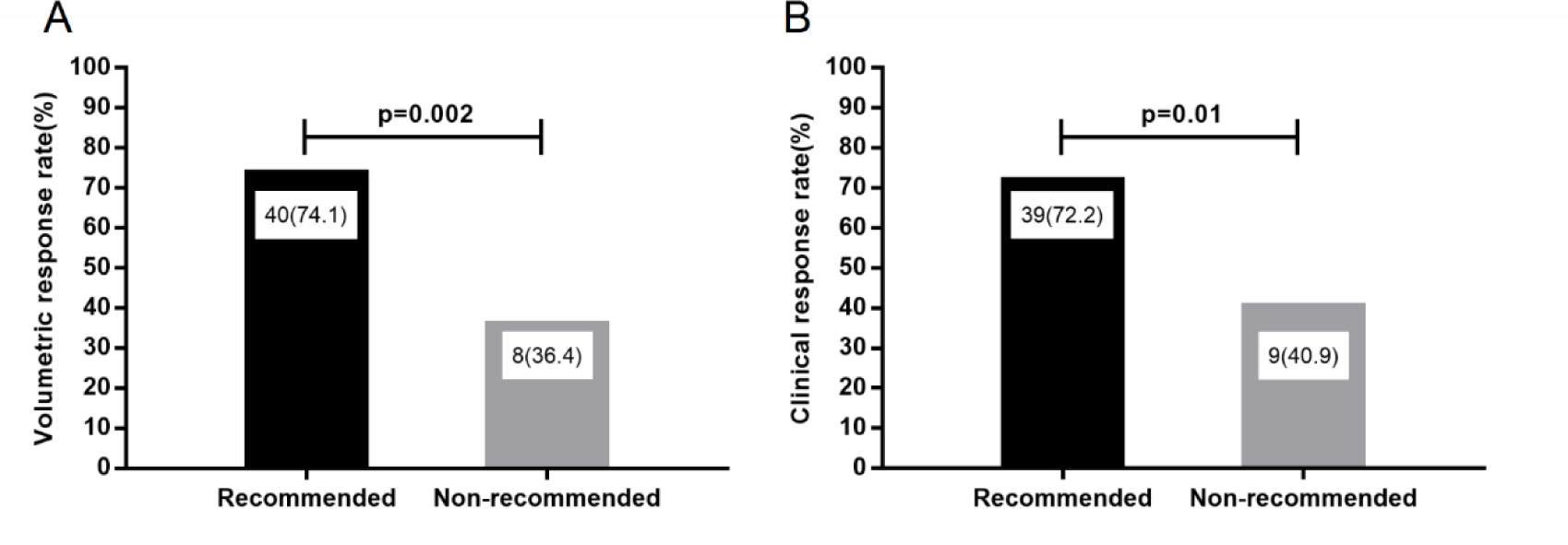
CRT response rates in the recommended and non-recommended groups. **(A)** CRT volumetric response rates in patients with LV leads in the recommended segment (n = 54) and patients with LV leads in the non-recommended segment (n = 22) (74.1% vs. 36.4%, *P* = .002). **(B)** CRT clinical response rates in patients with LV leads in the recommended segment (n = 54) and patients whose LV leads were in the non-recommended segments (n = 22) (72.2% vs. 40.9%, *P* = .01).

We validated our recommendation method in subgroup analyses. The actual LV lead was located in the lateral wall in 62 patients and non-lateral in 14 patients, the response rate showed no significance difference between the two groups (67.7% vs. 42.9%, *p*=.081) (Figure 4A). Further analysis of patients with a LV lead located in the lateral wall found that the response rate was 77.6% (38 of 49) in patients with a LV lead located in the recommended segment, whereas 30.8% (4 of 13) in patients with a non-recommended LV lead position (*p*= .004) (Figure 4B). Similar results were found in patients with QRSd ≥150ms and in patients with LBBB morphology; the response rate was significantly higher in patients with a LV lead located in the recommended segment than in patients with a non-recommended LV lead position (83.0% vs. 47.1%, *p* =.011; 82.6% vs. 50.0%, *p* =.025) (Figure 4C and 4D).

**Figure 4.**
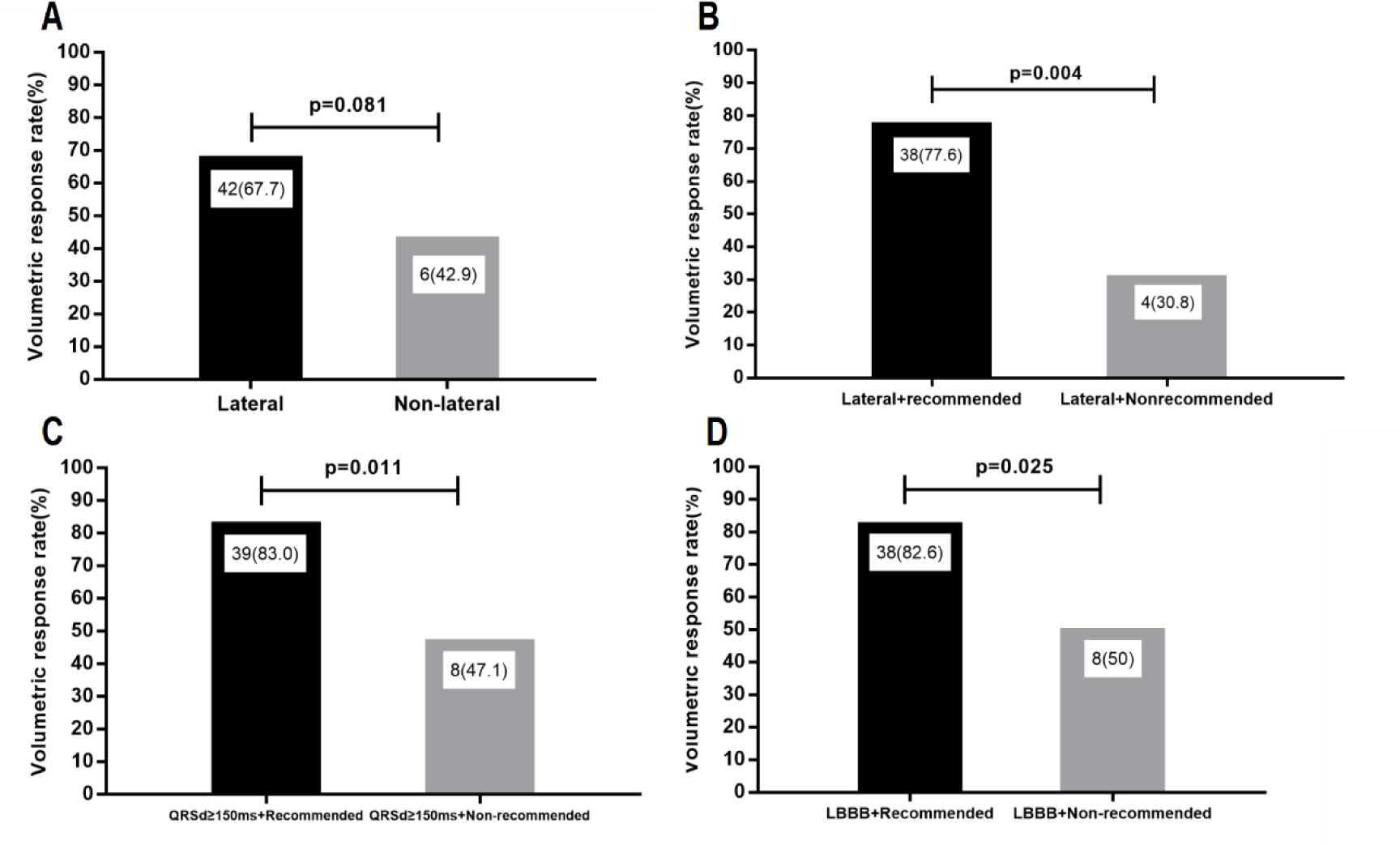
Subgroup analyses of CRT volumetric response. **(A)** CRT volumetric response rates in patients with LV leads in the lateral segment (n = 62) and patients with LV leads in the non-lateral segments (n = 14) (67.7% vs. 42.9%, *P* = .081). **(B)** CRT volumetric response rates in patients with LV leads in the lateral segment and meeting our recommended criteria (n=49) and patients with LV leads in the lateral segment and not meeting our recommended criteria (n=13) (77.6% vs. 30.8%, *P* = .004). **(C)** CRT volumetric response rates in patients whose QRSd≥150ms and meeting our recommended criteria (n=47) and patients whose QRSd≥150ms and not meeting our recommended criteria (n=17) (83.0% vs. 47.1%, *P* = .011). **(D)** CRT volumetric responses rate in patients with LBBB morphology and meeting our recommended criteria (n=46) and patients with LBBB morphology and not meeting our recommended criteria (n=16) (82.6% vs. 50.0%, *P* = .025).

#### Clinical response

In the recommended group, NYHA functional class improved by ≥ 1 in 72.2% of patients (n = 39) compared with only 40.9% of patients (n = 9) in the non-recommended group (*p* = .01) (Figure 3B).

### Univariable and Multivariable Regression Analyses

Univariable and multivariable logistic regression analyses were used to determine which parameters predict the CRT volumetric response. In the univariate regression analysis, presence of diabetes, QRSd, LBBB morphology, scar burden, and recommended LV lead position were associated with the volumetric response. In multivariable regression analyses, LV lead placement in the recommended segments versus non-recommended position remained an independent predictor of the volumetric response (odds ratio 7.326; 95% confidence interval 1.527-35.144, *p* =.013) (Table 2).

**Table 2.** Univariate analysis and multivariable models for the CRT volumetric response BMI, Body Mass Index; QRSd, QRS duration; LBBB, left bundle branch block; ACEi, angiotensin-converting enzyme inhibitors; ARB, angiotensin receptor blocker; LVEDV, left ventricular end-diastolic volume; LVESV, left ventricular end-systolic volume; MR, mitral regurgitation; TR, tricuspid regurgitation; PSD, phase standard deviation; PBW, phase histogram bandwidth

|  | Univariate |  | Multivariate |  |
| --- | --- | --- | --- | --- |
|  | OR(95% CI) | pvalue | OR(95% CI) | pvalue |
| Age | 0.985(0.947-1.024) | .439 |  |  |
| Male | 0.652(0.203-2.095) | .473 |  |  |
| BMI | 0.967(0.863-1.083) | .561 |  |  |
| Hypertension | 0.976(0.383-2.487) | .96 |  |  |
| Diabetes | 0.257(0.081-0.814) | .021 | 0.392(0.075-2.049) | .267 |
| Smoking | 1.202(0.465-3.105) | .704 |  |  |
| Lateral | 2.800(0.856-9.157) | .089 |  |  |
| QRSd | 1.043(1.016-1.071) | .002 | 1.048(1.008-1.089) | .018 |
| LBBB | 17.250(3.478-85.559) | <.0001 | 5.156(0.773-34.392) | .090 |
| Ischemic etiology | 2.391(0.585-9.777) | .225 |  |  |
| ACEi/ARB(%) | 0.633(0.178-2.249) | .48 |  |  |
| Beta-blocker(%) | 0.259(0.030-2.274) | .223 |  |  |
| Aldosterone antagonist(%) | 0.600(0.145-2.477) | .48 |  |  |
| Digoxin(%) | 0.487(0.166-1.427) | .190 |  |  |
| LVEDV(ml) | 0.995(0.989-1.000) | .066 |  |  |
| LVESV(ml) | 0.994(0.987-1.000) | .050 |  |  |
| Moderate/severe MR(%) | 0.292(0.059-1.445) | .131 |  |  |
| Moderate/severe TR(%) | 0.444(0.165-1.200) | .110 |  |  |
| PSD | 0.990(0.966-1.016) | .457 |  |  |
| PBW | 0.999(0.993-1.005) | .821 |  |  |
| Scar burden% | 0.930(0.885-0.977) | .004 | 0.901(0.842-0.965) | .003 |
| Wall thickening | 0.994(0.960-1.030) | .751 |  |  |
| Recommended vs. non-recommended | 5.000(1.731-14.441) | .003 | 7.326(1.527-35.144) | .013 |
| VCG coordination | 0.429(0.128-1.437) | .170 |  |  |
| SPECT coordination | 1.615(0.632-4.129) | .317 |  |  |

## DISCUSSION

In this study, we employed a two-pronged approach to determine the optimal LV lead placement in CRT patients. We identified the LMC segments through SPECT MPI phase analysis and the LEA site by VCG. The integration of these two techniques yielded promising results and CRT patients who received LV lead in the recommended position demonstrated a significantly higher volumetric response. Furthermore, our multivariable regression analyses strengthened this association, indicating that LV lead placement in the recommended segments was an independent predictor of the volumetric response after 6 months of CRT.

### Attempts for Improved CRT Response

LV lead position is known to be an important factor in determining response to CRT. Current practice for CRT is to place the LV lead at the lateral or posterolateral wall based on the benefit shown in early hemodynamic studies and the observation that the latest activated segment predominates at these sites. However, it has been controversial to direct the LV lead to the lateral or posterolateral wall. The MADIT-CRT study demonstrated that the benefit from CRT was similar for LV leads positioned along the anterior, lateral, or posterior wall.^5^ In the present study, we also found that the response rate showed no significant difference in patients with a LV lead located on the lateral wall compared with on the non-lateral wall (67.7% vs. 42.9%, *p*=.081). In addition, we found a volumetric response rate of only 30.8% in patients with a LV lead located on the lateral wall while the lateral wall was not our recommended position, further suggesting that the lateral wall may not be the optimal location for some patients.

QRS duration and morphology are used to select candidates for CRT. It has a class IA indication for symptomatic heart failure patients with sinus rhythm and a QRSd≥150 ms and LBBB morphology and with LVEF≤35% despite optimized medical treatments according to the 2021 ESC Guidelines on CRT.^22^ Several studies showed that patients with LBBB and prolonged QRS duration (QRSd≥150 ms) were more likely to be echocardiographic responders and were associated with favorable clinical outcomes.^23,24^ In subgroup analysis of the current study, 62 patients had LBBB morphology, 64 patients had a QRSd ≥150ms, and 57 patients had LBBB morphology with QRSd ≥150ms. We found that in patients with LBBB morphology, the response rate was higher when the LV lead was placed in the recommended position than when it was placed in the non-recommended position (82.6% vs. 50.0%, *p* =.025). The same result was found in patients with QRSd≥150ms (83.0% vs. 47.1%, *p* =.011). Furthermore, in patients with LBBB morphology and QRSd ≥150ms, the response rate was 88.1% (37 of 42) when the LV lead was placed in the recommended position, and was significantly higher than that when placed in the non-recommended position (88.1% vs. 53.3%, *p*=.014). This seems that our recommended LV lead position could further improve CRT response after strict guideline-directed patient selection.

A previous study demonstrated that implanting LV lead in the scarred region had a detrimental impact on LV reverse remodeling and increased clinical events.^9^ In the present study, we excluded scarred segments and found overall myocardial scar burden quantified by SPECT MPI predicted the volumetric response to CRT, suggesting that reverse remodeling requires viable myocardium adjacent to the LV lead.

### The latest mechanical contraction segment and electrical activation site

Several clinical studies have claimed that LV lead placement directs toward the segment of LMC improves the CRT response, including the TARGET,^6^ the STARTER,^25^ and the Imaging CRT study.^26^ These studies employed various imaging modalities, such as speckle tracking echocardiography, cardiac computed tomography (CT) and SPECT MPI, to guide the LV lead placement toward the LMC segment. Pacing remote from the LEA site may diminish the CRT response, as supported by studies utilizing non-invasive three-dimensional electrical activation mapping, a technique used to assess the distance of pacing site and the position of the LEA site. It was found that LV pacing close to the site of LEA provides improved volumetric response to CRT.^11^ VCG offers a more comprehensive and accurate assessment of cardiac electrical activity by capturing both the direction and magnitude of the cardiac electrical vectors. VCG can be used to measure the LEA site of the heart. The segment of LMC does not always overlap with the site of LEA. In this study, we advocate a novel approach for LV lead placement, integrating information from the segment with the LMC and the site with the LEA. Our findings indicate that pacing in the recommended LV lead segments is significantly associated with an increased CRT volumetric response.

Although the recent studies of novel CRT by conduction system pacing showed better efficacy of cardiac function and clinical outcomes compared to the traditional biventricular pacing,^27,28,29^ most of the enrolled patients were class one CRT recommendation with typical LBBB. To our best knowledge, CRT delivered by conduction system pacing was referred as at most IIa level recommendation at present.^30^ This new integration of LMC and LEA may be very useful to improve traditional biventricular pacing CRT, especially for non-LBBB patients.

### Limitations

This retrospective observational study had a limited sample size, and data collection occurred between 2011 and 2021. It’s worth noting that the guidelines for CRT have evolved, and optimal medical therapy has incorporated additional drug classes. Subgroup analyses were conducted, stratified by ECG morphology and QRS duration; however, caution is advised in interpreting the results due to the relatively low number of non-LBBB patients and those with QRS duration <150ms in this cohort. Larger prospective multicenter studies are warranted to provide more robust insights.

## CONCLUSIONS

Pacing in the LV lead segments recommended by the integration of both mechanical and electrical dyssynchrony was associated with an increased CRT volumetric response. The findings from this study underscore the importance of conducting larger and prospective studies in the future to further validate these results.

## Data Availability

All data produced in the present study are available upon reasonable request to the authors

## Acknowledgements

This research was supported by National Natural Science Foundation of China (82070521), Clinical Competence Improvement Project of Jiangsu Province Hospital (JSPH-MA-2020-3, JSPH-MA-2022-2, JSPH-MB-2022-11), and a Project on New Technology of Jiangsu Province (JX233C202103)

## References

1. Vernooy K, van Deursen CJ, Strik M, Prinzen FW. Strategies to improve cardiac resynchronization therapy. Nature reviews Cardiology 2014;11:481–93.

2. Daubert C, Behar N, Martins RP, Mabo P, Leclercq C. Avoiding non-responders to cardiac resynchronization therapy: a practical guide. European heart journal 2017;38:1463–72.

3. Gold MR, Birgersdotter-Green U, Singh JP, Ellenbogen KA, Yu Y, Meyer TE et al. The relationship between ventricular electrical delay and left ventricular remodelling with cardiac resynchronization therapy. European heart journal 2011;32:2516–24.

4. Kydd AC, Khan FZ, Watson WD, Pugh PJ, Virdee MS, Dutka DP. Prognostic benefit of optimum left ventricular lead position in cardiac resynchronization therapy: follow-up of the TARGET Study Cohort (Targeted Left Ventricular Lead Placement to guide Cardiac Resynchronization Therapy). JACC Heart failure 2014;2:205–12.

5. Singh JP, Klein HU, Huang DT, Reek S, Kuniss M, Quesada A et al. Left ventricular lead position and clinical outcome in the multicenter automatic defibrillator implantation trial-cardiac resynchronization therapy (MADIT-CRT) trial. Circulation 2011;123:1159–66.

6. Khan FZ, Virdee MS, Palmer CR, Pugh PJ, O’Halloran D, Elsik M et al. Targeted left ventricular lead placement to guide cardiac resynchronization therapy: the TARGET study: a randomized, controlled trial. Journal of the American College of Cardiology 2012;59:1509–18.

7. Zhou W, Garcia EV. Nuclear Image-Guided Approaches for Cardiac Resynchronization Therapy (CRT). Current cardiology reports 2016;18:7.

8. Zou J, Hua W, Su Y, Xu G, Zhen L, Liang Y et al. SPECT-Guided LV Lead Placement for Incremental CRT Efficacy: Validated by a Prospective, Randomized, Controlled Study. JACC Cardiovascular imaging 2019;12:2580–3.

9. Zhang X, Qian Z, Tang H, Hua W, Su Y, Xu G et al. A new method to recommend left ventricular lead positions for improved CRT volumetric response and long-term prognosis. Journal of nuclear cardiology : official publication of the American Society of Nuclear Cardiology 2021;28:672–84.

10. Mafi Rad M, Blaauw Y, Dinh T, Pison L, Crijns HJ, Prinzen FW et al. Different regions of latest electrical activation during left bundle-branch block and right ventricular pacing in cardiac resynchronization therapy patients determined by coronary venous electro-anatomic mapping. European journal of heart failure 2014;16:1214–22.

11. Parreira L, Tsyganov A, Artyukhina E, Vernooy K, Tondo C, Adragao P et al. Non-invasive three-dimensional electrical activation mapping to predict cardiac resynchronization therapy response: site of latest left ventricular activation relative to pacing site. Europace : European pacing, arrhythmias, and cardiac electrophysiology : journal of the working groups on cardiac pacing, arrhythmias, and cardiac cellular electrophysiology of the European Society of Cardiology 2023;25:1458–66.

12. Mafi Rad M, Wijntjens GW, Engels EB, Blaauw Y, Luermans JG, Pison L et al. Vectorcardiographic QRS area identifies delayed left ventricular lateral wall activation determined by electroanatomic mapping in candidates for cardiac resynchronization therapy. Heart rhythm 2016;13:217–25.

13. Nguyên UC, Claridge S, Vernooy K, Engels EB, Razavi R, Rinaldi CA et al. Relationship between vectorcardiographic QRS(area), myocardial scar quantification, and response to cardiac resynchronization therapy. Journal of electrocardiology 2018;51:457–63.

14. Man S, Maan AC, Schalij MJ, Swenne CA. Vectorcardiographic diagnostic & prognostic information derived from the 12-lead electrocardiogram: Historical review and clinical perspective. Journal of electrocardiology 2015;48:463–75.

15. Wang J, Wang Y, Yang M, Shao S, Tian Y, Shao X et al. Mechanical contraction to guide CRT left-ventricular lead placement instead of electrical activation in myocardial infarction with left ventricular dysfunction: An experimental study based on non-invasive gated myocardial perfusion imaging and invasive electroanatomic mapping. Journal of nuclear cardiology : official publication of the American Society of Nuclear Cardiology 2020;27:419–30.

16. van der Bijl P, Kostyukevich MV, Khidir M, Ajmone Marsan N, Delgado V, Bax JJ. Left ventricular remodelling and change in left ventricular global longitudinal strain after cardiac resynchronization therapy: prognostic implications. European heart journal Cardiovascular Imaging 2019;20:1112–9.

17. Lin X, Xu H, Zhao X, Chen J. Sites of latest mechanical activation as assessed by SPECT myocardial perfusion imaging in ischemic and dilated cardiomyopathy patients with LBBB. European journal of nuclear medicine and molecular imaging 2014;41:1232–9.

18. Boogers MJ, Chen J, van Bommel RJ, Borleffs CJ, Dibbets-Schneider P, van der Hiel B et al. Optimal left ventricular lead position assessed with phase analysis on gated myocardial perfusion SPECT. European journal of nuclear medicine and molecular imaging 2011;38:230–8.

19. Wang CY, Hung GU, Lo HC, Tsai SC, He Z, Zhang X et al. Clinical impacts of scar reduction on gated myocardial perfusion SPECT after cardiac resynchronization therapy. Journal of nuclear cardiology : official publication of the American Society of Nuclear Cardiology 2022;29:2571–9.

20. Jaros R, Martinek R, Danys L. Comparison of Different Electrocardiography with Vectorcardiography Transformations. Sensors (Basel, Switzerland) 2019;19.

21. Zou F, Brar V, Worley SJ. Interventional device implantation, Part I: Basic techniques to avoid complications: A hands-on approach. Journal of cardiovascular electrophysiology 2021;32:523–32.

22. Glikson M, Nielsen JC, Kronborg MB, Michowitz Y, Auricchio A, Barbash IM et al. 2021 ESC Guidelines on cardiac pacing and cardiac resynchronization therapy. European heart journal 2021;42:3427–520.

23. Oka T, Inoue K, Tanaka K, Hirao Y, Isshiki T, Kimura T et al. Effect of QRS Morphology and Duration on Clinical Outcomes After Cardiac Resynchronization Therapy - Analysis of Japanese Multicenter Registry. Circulation journal : official journal of the Japanese Circulation Society 2018;82:1813–21.

24. van der Bijl P, Khidir M, Leung M, Mertens B, Ajmone Marsan N, Delgado V et al. Impact of QRS complex duration and morphology on left ventricular reverse remodelling and left ventricular function improvement after cardiac resynchronization therapy. European journal of heart failure 2017;19:1145–51.

25. Saba S, Marek J, Schwartzman D, Jain S, Adelstein E, White P et al. Echocardiography-guided left ventricular lead placement for cardiac resynchronization therapy: results of the Speckle Tracking Assisted Resynchronization Therapy for Electrode Region trial. Circulation Heart failure 2013;6:427–34.

26. Sommer A, Kronborg MB, Nørgaard BL, Poulsen SH, Bouchelouche K, Böttcher M et al. Multimodality imaging-guided left ventricular lead placement in cardiac resynchronization therapy: a randomized controlled trial. European journal of heart failure 2016;18:1365–74.

27. Wang Y, Zhu H, Hou X, Wang Z, Zou F, Qian Z et al. Randomized Trial of Left Bundle Branch vs Biventricular Pacing for Cardiac Resynchronization Therapy. Journal of the American College of Cardiology 2022;80:1205–16.

28. Diaz JC, Sauer WH, Duque M, Koplan BA, Braunstein ED, Marín JE et al. Left Bundle Branch Area Pacing Versus Biventricular Pacing as Initial Strategy for Cardiac Resynchronization. JACC Clinical electrophysiology 2023;9:1568–81.

29. Vijayaraman P, Sharma PS, Cano Ó, Ponnusamy SS, Herweg B, Zanon F et al. Comparison of Left Bundle Branch Area Pacing and Biventricular Pacing in Candidates for Resynchronization Therapy. Journal of the American College of Cardiology 2023;82:228–41.

30. Chung MK, Patton KK, Lau CP, Dal Forno ARJ, Al-Khatib SM, Arora V et al. 2023 HRS/APHRS/LAHRS guideline on cardiac physiologic pacing for the avoidance and mitigation of heart failure. Heart rhythm 2023;20:e17–e91.

